# Opportunities for more powerful statistical analyses of ordinal scales: A scoping review of neurological trials

**DOI:** 10.1101/2024.06.24.24309421

**Authors:** Yongxi Long, Sophie C. de Ruiter, Linda W.G. Luijten, Eveline J.A. Wiegers, Diederik W.J. Dippel, Pieter A. van Doorn, Bart C. Jacobs, Ewout W. Steyerberg

## Abstract

**Background and Objectives:** Ordinal scales are widely adopted as outcome measures in neurological randomized controlled trials (RCTs). There have been extensive discussions about appropriate statistical analysis strategies of ordinal neurological outcomes. We aimed to evaluate which statistical methods have been used to test and estimate treatment effects from ordinal outcomes in recent RCTs across a range of acute neurological diseases.

**Methods:** We searched for RCTs in five acute monophasic neurological diseases (stroke, traumatic brain injury (TBI), subarachnoid hemorrhage (SAH), meningitis, and Guillain-Barré Syndrome (GBS)) published in high-impact journals between January 1, 2015 and November 1, 2023. Trials had to report on an ordinal scale as the primary or secondary outcome. Two independent reviewers assessed whether/how investigators (1) delt with the ordinal nature of outcomes, (2) assessed and reported key assumptions,(3)utilized longitudinal measurements, (4) adjusted for prognostic variables.

**Results:** We included 70 RCTs for treatment evaluations in stroke (n=36), TBI (n=13), SAH (n=10), meningitis (n=7), and GBS (n=4). In 46/70 (66%) trials, investigators retained the full ordering information, commonly analyzed by a proportional odds model (33/46 trials, 72%). The proportional odds assumption was not addressed in 23/33 (62%) of these trials. In 22/70 (31%) trials, the ordinal outcome was dichotomized, with notable disagreement on the cut-point within neurological diseases. In 41/70 (59%) trials, the ordinal outcome was assessed at multiple time points, while some form of longitudinal data analysis was performed in only three (7%) of these 41 studies. The time point chosen for analysis was inconsistent within neurological diseases.

**Discussion:** The current practice of analyzing ordinal outcomes is often suboptimal in neurological trials according to modern statistical standards. Dichotomization and focus on a single arbitrary time point are still common, while more efficient analysis strategies exist. Further research needs to clarify the balance between maximizing the statistical power and assumptions made in approaches that better leverage ordinal information.

## INTRODUCTION

Randomized controlled trials (RCTs) play a crucial role in providing evidence for the effectiveness of novel treatments to inform healthcare decisions. For example, a series of RCTs has led to the major breakthrough of establishing endovascular therapy as a key component of acute ischemic stroke care (Saber et al., 2019). However, in other acute monophasic neurological diseases, such as Guillain-Barré Syndrome (GBS) and traumatic brain injury (TBI), resource limitations and substantial disease heterogeneity pose challenges to the conduct of RCTs. Over the last 30 years, most RCTs for new therapies in GBS reported neutral results (Leonhard et al., 2019).

Outcome analysis forms an important aspect of treatment investigations. Ordinal scoring scales such as the modified Ranking Scale (mRS) (Banks & Marotta, 2007) have been widely used for outcome assessment in neurological trials. However, a common practice is to dichotomize the ordinal scale into favorable versus unfavorable conditions. Dichotomization loses information on the full range of possible statuses and suffers from a loss of power (van Leeuwen et al., 2019). Moreover, dichotomization forces patients with different clinical features to cross the same threshold defined by an arbitrary cut-point to contribute to a positive treatment effect. For patients with poor baseline prognosis profiles, shifts towards better conditions while still below the cut-point value may still be clinically meaningful.

Therefore, it is desirable to exploit the rank ordering information of ordinal outcomes to increase statistical power. One common ordinal analysis approach is the proportional odds model. Ordinal analysis is particularly appealing for trials in rare neurological diseases since it is often infeasible to reach the desired power with a dichotomous outcome design due to the scarcity of patients(D’Amico et al., 2020; Ganesh et al., 2018). Many trials had repeated outcome measurements on the same patients over time. Aside from considering ordinal analysis at a predetermined fixed time point, longitudinal analysis incorporating information from repeated measurements may provide additional statistical power. Especially in heterogeneous diseases like TBI as it can be challenging to decide which time point is the most representative of the treatment effect on clinical recovery (Tang et al., 2018).

We aimed to evaluate recent practice of analyzing ordinal outcomes in neurological trials, with a focus on five acute monophasic neurological diseases (stroke, GBS, TBI, meningitis, and subarachnoid hemorrhage (SAH)). Summarizing the current statistical practice would highlight opportunities for more efficient design and analysis for future neurological trials.

## METHODS

### Search strategy

We searched PubMed for recent RCTs in stroke, GBS, TBI, meningitis, and SAH, that were published in top neurological journals between January 1, 2016 and May 1, 2021. Search terms are presented in Appendix 1. We initially limited the total number to 50 trials for feasibility. We prioritized inclusion of all trials from the four less prevalent diseases, then we allocated the remaining slots to the most recent stroke trials. In November 2023, we updated the search to include trials until November 1, 2023 and expanded to 70 trials.

### Inclusion/exclusion criteria

#### The review included RCTs that met the following criteria

1. The RCT had to report on at least one ordinal scale as the primary or secondary efficacy outcome in one of the five neurological diseases of interest.
2. The RCT had to be published in a journal with an impact factor of at least five in the year 2021 (relaxed to an impact factor of at least two for GBS as a rare disease).

We excluded studies that started the intervention after the acute phase of the disease. These studies mostly focused on comorbidity or rehabilitation (e.g., depression after stroke).

#### Screening and data extraction

Eligible trial reports were imported to Covidence for screening and extraction (Covidence systematic review software, Veritas Health Innovation, Melbourne, Australia). If information was not clear in the trial report, we further referred to trial protocols and supplementary materials. Two independent reviewers assessed whether ordinal outcomes were dichotomized, analyzed as ordinal, or as continuous variables. We considered outcomes with 15 or more possible values as being continuous, such as the Barthel Index (Song et al., 2006; Walters et al., 2001). When more than one method was used for analyzing the ordinal outcome, we considered the statistically best alternative. For example, we scored a trial as “ordinality exploited” when the ordinal scale was both analyzed using a Mann-Whitney U test and a binary logistic regression model. We recorded the number of measurements collected over time and assessed whether a longitudinal analysis was done. In addition, we evaluated whether covariate adjustment and subgroup analysis were performed. Appendix 2 provides items that were extracted.

#### Statistical analysis

We considered the following statistical methods as approaches for exploiting ordinality:

1. The proportional odds (PO) model, also known as the “shift analysis”, providing a summary measure of the treatment effect by estimating a common odds ratio over all possible cut-points of the outcome scale(McCullagh, 1980; Valenta et al., 2006). The PO model assumes that the association between each predictor with the ordinal outcome is identical across these cut-points, a condition known as the *Proportional Odds Assumption*.
2. The Mann-Whitney U test (also called Wilcoxon rank sum test) and its associated effect size measures such as the win odds(Churilov et al., 2022). The win odds represents the odds of “winning”, i.e. a randomly sampled patient from the treated group achieving a more favorable level at the ordinal outcome scale than one from the control group (with ties evenly split).
3. The t-test, which was shown to be valid for ordinal data(Heeren & D’Agostino, 1987).

RCTs often have multiple secondary outcomes in addition to their primary outcomes. We reasoned that different analysis methods might be adopted under different research goals. Therefore, we stratified primary and secondary ordinal outcomes. For primary ordinal outcomes, we further distinguished whether ordinality was exploited in the main analysis, in the supplementary analysis (including secondary, exploratory, sensitivity, and post-hoc analysis), or not exploited. For secondary ordinal outcomes, we only examined whether ordinality was exploited.

To explore the factors influencing the utilization of ordinal analyses (yes/no), we performed a multivariable logistic regression considering publication year, disease, and trial sample size.

## RESULTS

We included four RCTs in GBS, seven in meningitis, 10 in SAH, 13 in TBI and 36 in stroke (including 32 investigating ischemic stroke patients). One stroke RCT investigated intracerebral hemorrhage patients(Meretoja et al., 2020) and three stroke RCTs had a mixed population of ischemic stroke and intracerebral hemorrhage patients(Bösel et al., 2022; Hankey et al., 2020; Lundström et al., 2020) (Appendix 3 provides details for extracted information).

### Data extraction and reviewer agreement

The two independent reviewers agreed on the majority of the papers (90%, SR and LL for the initial 50 papers, YL and LL for updated 20 papers). For six papers (8%) there were small discrepancies, which were resolved after discussion. Analysis of one paper (1%) remained unclear, but after discussion with a third reviewer (ES), all discrepancies were resolved.

### Ordinal analysis

Measures that were commonly used for evaluating neurological functionality were the GBS disability scale (GBS-DS) for GBS, mRS for meningitis, SAH and stroke, Glasgow Outcome Scale (GOS) for SAH, and Glasgow Outcome Scale-Extended (GOS-E) for SAH and TBI (detailed definitions presented in Appendix 1).

Among the 70 papers that were reviewed, 46 (66%) respected the ordinality when analyzing ordinal outcomes (Figure 1). In 22 trials (31%) the ordinal scale was dichotomized either by a cut-point or by whether there was a favorable improvement from baseline. The common threshold for improvement was one-point increase/decrease in the scores, but some studies evaluated a two-point improvement (Anetsberger et al., 2020; Heemskerk et al., 2016; Heemskerk et al., 2017; Wong et al., 2015). Two studies treated the ordinal scale as continuous and performed mean comparison by a t-test.

**Figure 1.**
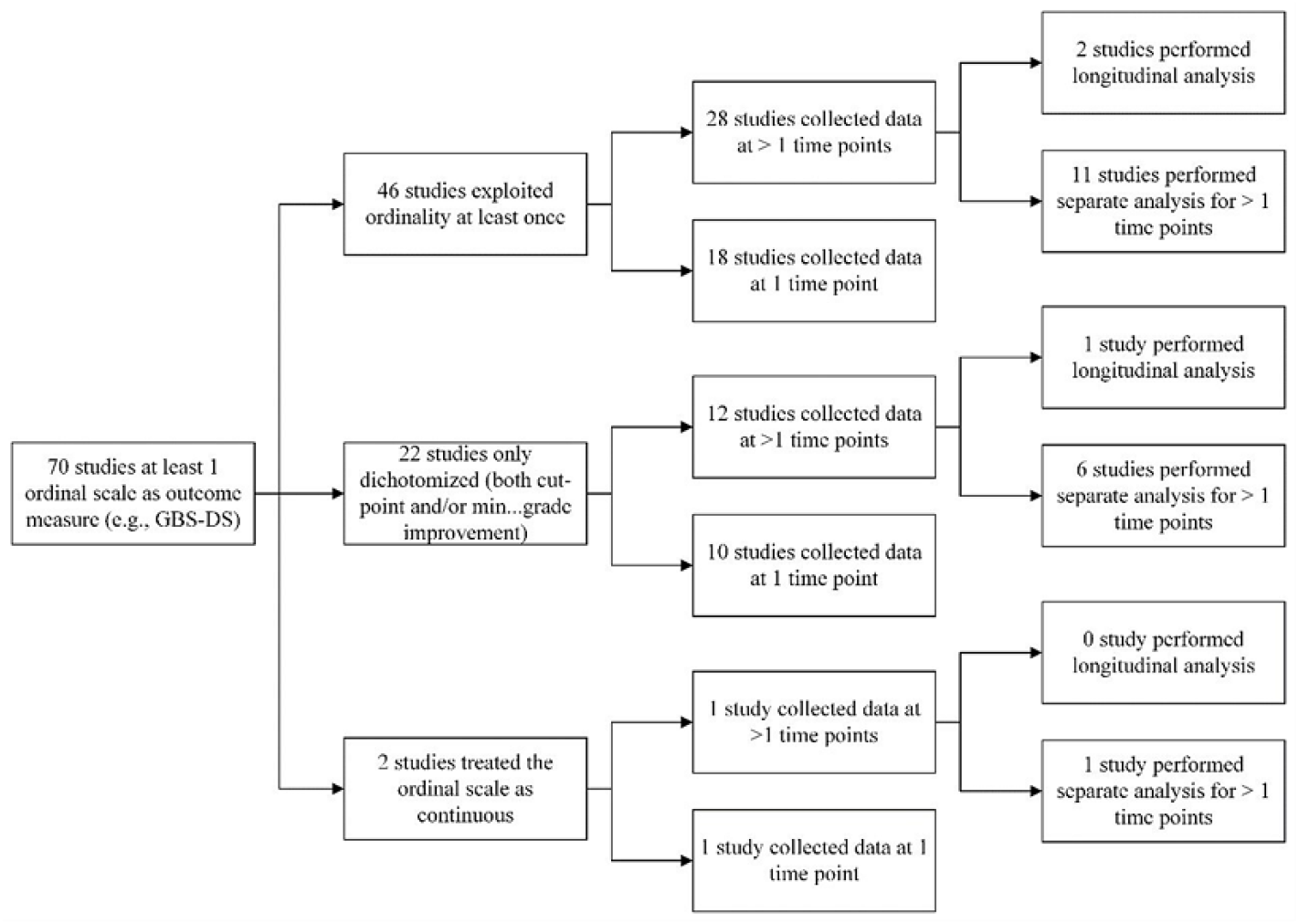
Flowchart on the data collection and analysis of ordinal outcomes of 70 included studies. For studies that collected data at > 1 time points, some performed a single time point analysis only.

Stroke and TBI trials were more likely to adopt ordinal analyses compared to trials in other fields (odds ratio 5.6, 95% CI (1.4, 24); and 3.5, 95% CI (0.8, 18) respectively, Table 1).

**Table 1.** Descriptives of included studies and relation with ordinal analysis (multivariable logistic regression analysis).

| Factor | Ordinal analysis | Odds ratio (95% CI) |
| --- | --- | --- |

|  | Yes (n=46) | No (n=24) |  |
| --- | --- | --- | --- |
| **Year, mean (sd)** | 2020 (2.4) | 2019 (2.4) | 1.1 (0.8, 1.3) |
| **Field, n (%)<sup>\*</sup>** |  |  |  |
| GBS | 1 (2) | 3 (13) |  |
| Meningitis | 4 (9) | 3 (13) | 1.0 (Reference)<sup>¶</sup> |
| SAH | 3 (7) | 7 (29) |  |
| TBI | 9 (20) | 4 (17) | 3.5 (0.8, 18) |
| Stroke | 29 (63) | 7 (29) | 5.6 (1.4, 24) |
| **Sample size, median (IQR)** | 326 (192-569) | 99 (51-579) | 1.1 (0.7, 1.7)<sup>§</sup> |

\* Percentage may not add up to 100% due to rounding
¶ For the reference category we used the pooled group including trials for GBS, meningitis and SAH, because of the low number of trials in these fields.
§ Sample size is right skewed so log transformation was performed in the multivariable logistic regression analysis

Within the 70 included RCTs a total of 39 ordinal outcomes were assessed as primary outcomes, and 64 as secondary (details in Appendix 3). Most primary ordinal outcomes (74%) were analyzed using methods that exploit ordinality, either as a main analysis (48%) or a supplementary analysis (52%) (Appendix 1, Figure 1A). PO models emerged as the prevailing method employed in both main and supplementary analyses. Nevertheless, 26% of primary ordinal outcomes were collapsed into a binary scale or treated as unordered. For secondary ordinal outcomes, more than half (56%) were dichotomized or treated as unordered. The Mann-Whitney U test, along with its associated effect size measures, was more common in the analysis of secondary ordinal outcomes (54%) compared to its utilization in the analysis of primary ordinal outcomes (14% in the main analysis and 7% in the supplementary analysis).

If a dichotomous analysis was performed, notable discrepancies regarding the cut-point were observed (Figure 2, see Table 1A. in Appendix 1 for tabular representation). The cut-points used for mRS varied considerably with the 2 to 3 threshold reported most often, followed by the 1 to 3 and 3 to 4 thresholds.

**Figure 2.**
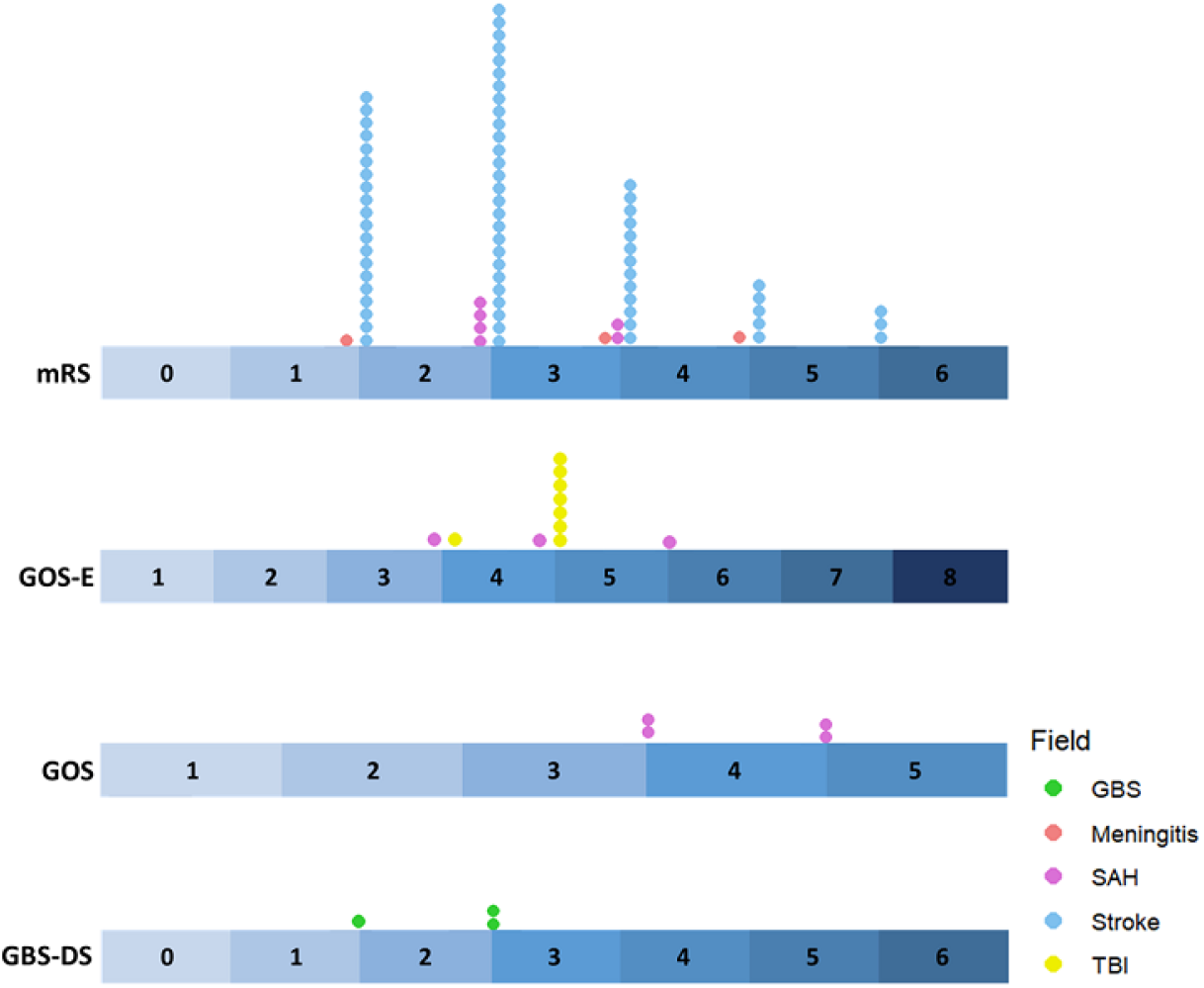
Frequency of cut-points chosen for dichotomization (shown by the number of dots above each ordinal scale) for common ordinal scales in GBS, Meningitis, SAH, stroke and TBI.

### Proportional odds assumption

Among 46 trials that performed at least one ordinal analysis, 33 (72%) adopted the proportional odds (PO) model method. A total of 37 PO models were fitted, with four studies reporting more than one PO model for multiple ordinal outcomes). Results from 23 (62%) PO models were reported without mentioning the assessment of the proportional odds assumption. Two of these studies mentioned testing for proportionality in their statistical analysis plan.. One study (Andrews et al., 2015) stated that if the proportional odds assumption is violated, treatment heterogeneity will be explored descriptively. The other study(Yang et al., 2022) wrote that they would proceed with the proportional odds analysis if the proportional assumption is violated but interpret it as a shift analysis. Results from 12 PO models were presented with explicit statements about the satisfaction of the proportional odds assumption, either by a Brant’s test, a likelihood ratio test, or a graphical check (Chabriat et al., 2020). Two PO model analyses were reported while the proportional odds assumption was violated after adjusting for baseline covariates (Martins et al., 2020; Post et al., 2021). Specifically, one study (Martins et al., 2020) examined odds ratios from separate binary analyses and found them all showing a positive treatment effect.

Six PO model analyses were originally planned but substituted by alternative methods after the violation of the proportional odds assumption. The non-parametric win odds method was adopted for three analyses (Campbell et al., 2020; Huo et al., 2023; Meretoja et al., 2020) and the unordered chi-square test was used for two analyses (Hutchinson et al., 2016). Another study applied dichotomization to the ordinal outcome(Hill et al., 2020).

### Repeated measurements and longitudinal analysis

Among the 46 trials that respected ordinality, 28 (61%) measured the ordinal outcome at more than one time point (Figure 1). Most studies performed analysis for only one of these time points (n=15, 54%) or performed multiple separate analyses for each of the time points (n=11, 39%). One trial analyzed the longitudinal dichotomized ordinal scale by generalized estimation equation (GEE) (Elkind et al., 2020). Another trial used a random quadratic model to classify patients into different clusters with respect to their speed of improvement on the ordinal outcome (Wu et al., 2023).

Disagreement about the time points used for analysis was observed for common ordinal outcome measures (Table 2). For mRS in stroke trials and GOS-E in TBI trials, there were relatively consistent choices: mRS at 90 days and GOS-E at 6 months respectively. In other cases, such as mRS in meningitis trials and SAH trials, ordinal measures were analyzed at a variety of different time points. Some studies also used time at discharge for analysis, which precluded a clear assessment of the absolute duration from baseline and varied per patient.

**Table 2.**
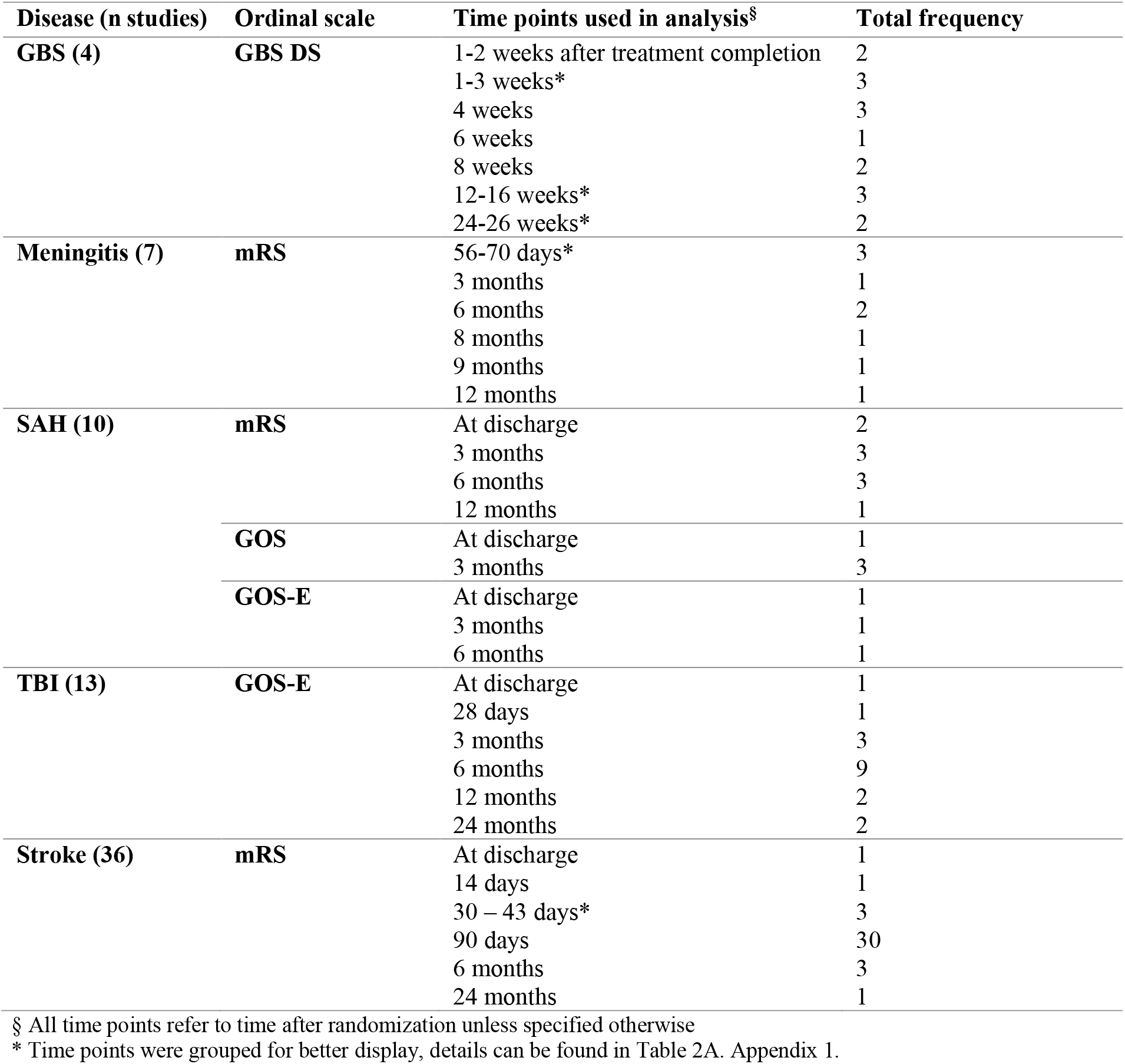
Frequency of time points selected for analysis for common ordinal measures in included 70 studies.

### Covariate adjustment and subgroup analysis

The majority of included trials (n=48, 69%) performed adjusted analysis by considering prognostic covariates, and more than half (n=38, 54%) performed subgroup analyses. Most studies pre-specified covariates to be adjusted for and subgroup analyses in their statistical analysis plans prior to the unblinding of the trial data. Several studies conducted post-hoc covariate adjustment as sensitivity analysis (Misawa et al., 2018) or post-hoc subgroup analysis (An et al., 2020; Pico et al., 2020). None of the trials adjusted for multiplicity (correction for p values when doing multiple testing) when performing subgroup analysis.

## DISCUSSION

This scoping review highlighted missed opportunities in the analysis of ordinal outcomes in recent neurological clinical trials. Dichotomization remained a common practice as about 30% of the included studies collapsed their ordinal outcomes and performed dichotomous analyses only. We observed non-consistent choices of cut-points for dichotomization, even for routinely used ordinal scales such as the mRS in stroke trials. There were various choices for the time point selected for analysis. The utilization of longitudinal methods for analyzing repeated ordinal measurements was uncommon.

We recognized that the choice of the cut-point could be influenced by specific conditions of the trial. For example, in a trial where a large proportion of poor outcomes were expected, the upper severe-disability category of GOS-E was included in the definition of favorable outcome (Hutchinson et al., 2016). Dichotomization usually results in a loss of statistical power to detect a treatment effect (Fedorov et al., 2009) and the choice on the cut-point can be arbitrary. Efficient analysis methods that retain the complete ranking of ordinal outcomes, such as the PO model, are desirable. We here provide a brief comparison of major statistical methods for analyzing ordinal outcomes including suggestions on when to consider each method (Table 3).

**Table 3.** Comparison of major statistical methods applicable to the analysis of ordinal scales.

| Ways to deal with ordinal scales | Example methods | Consider when | Do not consider when |
| --- | --- | --- | --- |
| **Dichotomizing (alive vs dead, favorable vs unfavorable)** | - Chi-squared test<br>- Binary logistic regression model | - There is an explicit clinical interest in contrasting a transition between two states | - The categories collapsed together are clinically rather different<br>- There is no clear cut-point |
| **Preserving the ordinal nature** | - Mann-Whitney U test and associated effect size measures<br>- Proportional odds model (ordered/ordinal logistic model, ordered/cumulative logit model/shift analysis) | - Ordinal categories are well-defined and transitions between different categories bear roughly similar clinical importance | - Transitions between ordinal categories are of hugely different clinical meaningfulness |
| **Treating as continuous** | - t-test<br>- Chi-squared test for trend<br>- ANOVA<br>- Linear regression | - Ordinal categories are well-defined and transitions between different categories bear roughly similar clinical importance<br>- Mean difference of the ordinal outcome can be sensibly interpreted | - Transitions between ordinal categories are of hugely different clinical meaningfulness. In this case, it is recommended to assign different numerical values (e.g., utility) to reflect such differences (Chaisinanunkul et al., 2015) |

There has been ongoing debate about the appropriateness of the PO model when the proportional odds assumption is violated. The Mann-Whitney U test is often seen as an alternative approach (Rahlfs et al., 2014). The PO model and the Mann-Whitney U test however are nearly equivalent in terms of testing the treatment effect. If a PO model gives a p value < 0.05, so does a Mann-Whitney U test (Wang & Tian, 2017). Of note, the common odds ratio (from the PO model) is less interpretable than effect size measures based on Mann-Whitney U test because it is built upon the proportional odds assumption. The common odds ratio from a PO model is a summary of the odds ratios of having a better outcome in the treatment arm compared to the control arm, assuming that the odds ratios are the same at each cut-point. Since the proportional odds assumption rarely holds exactly, the common odds ratio can merely be viewed as a weighted average of different odds ratios across all cut-points. The win odds (related to the Mann-Whitney U test) is the odds of a randomly sampled patient from the treatment arm having better outcomes than a randomly sampled patient from the control arm, without relying on the proportional odds assumption for interpretation (Churilov et al., 2022; Rahlfs et al., 2014)

There are statistical tests for assessing the proportional odds assumption, such as the Brant test and likelihood ratio test (Brant, 1990; Liu et al., 2023). Statistical significance does not necessarily imply a practical significance. Violations of proportionality can be either quantitative (different odds ratios at different cut-points, but all in the same direction) or qualitative (different odds ratios in the opposite directions, for example, the treatment yields more excellent outcomes at the cost of increased mortality). If the violation is quantitative and odds ratios for each dichotomization of the ordinal scale are reported for transparency, the common odds ratio may still provide an informative summary measure. When there is a qualitative violation of the proportionality, summarizing into a single effect is misleading no matter what ordinal analysis method is adopted. We then recommend to report descriptively, such as visualization by a Grotta chart.

For illustration, we discuss two large trials in neurology here. In the RESCUEicp trial, decompressive craniectomy was studied in TBI, and the outcome was measured by GOS-E (Hutchinson et al., 2016). The MR CLEAN trial investigated endovascular treatment in stroke, with the outcome measured by mRS (Berkhemer et al., 2014). Re-analysis is based on reconstructed datasets from original reports (see Appendix 4 for R code)

Grotta bars (Figure 3) provide an essential descriptive check for proportionality by visualizing the full distribution of the ordinal outcome. The PO assumption was qualitatively violated in the RESCUEicp trial, with a decrease in mortality (GOS-E=1), but an increase in the proportions of vegetative state (GOS-E=2) in the decompressive craniectomy group compared to the control group. Indeed, the odds ratio is negative for GOS-E 1-6 vs. 7 while positive for GOS-E 1 vs 2-7 (Appendix 1, Figure 2A). This means that patients in the decompressive craniectomy group were less likely to die but also less likely to achieve lower/upper good recovery compared to patients in the control group. This information would be hidden by a single summary measure. The original study reported results descriptively as differences between percentages. For the MR CLEAN trial, the treatment effect was positive across all cut-points. Therefore, it may be quite sensible to calculate a common odds ratio to represent a general beneficial shift towards better conditions.

**Figure 3.**
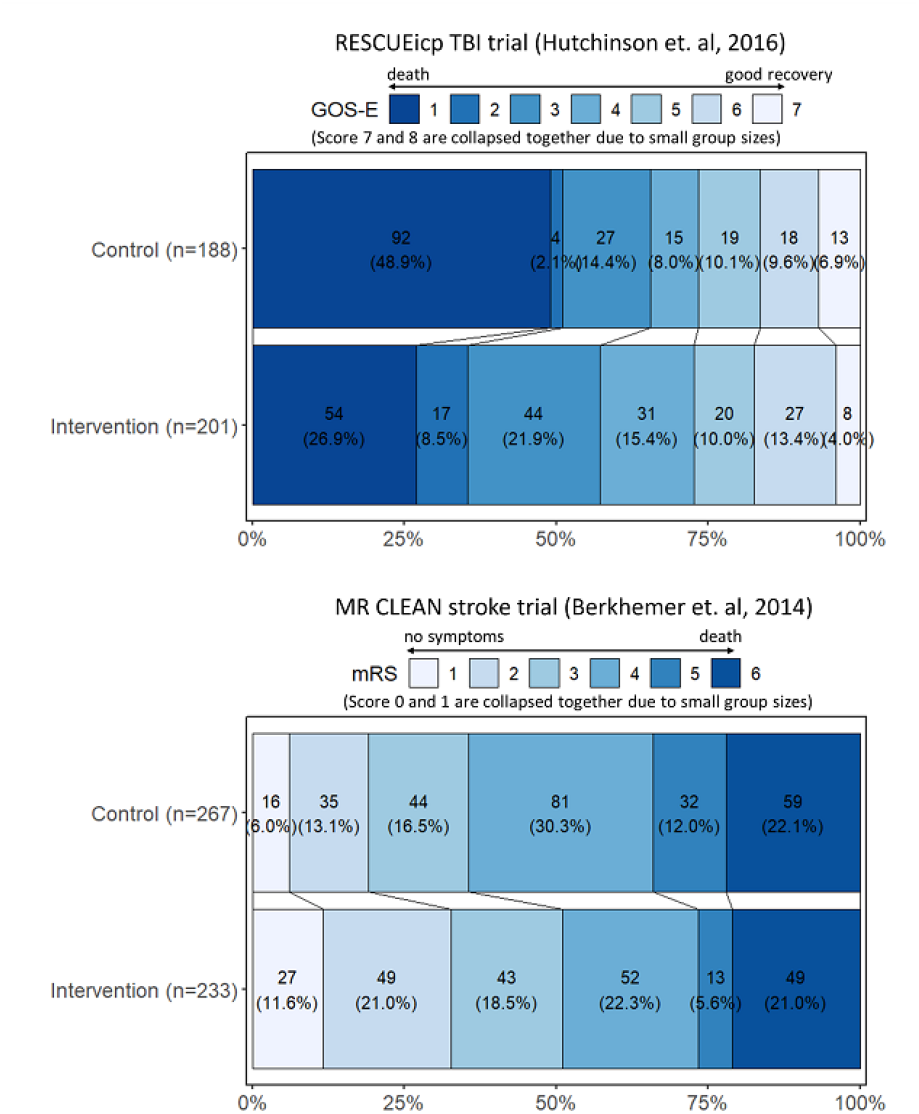
Grotta charts of the distribution of ordinal categories for RESCUEicp TBI trial (upper panel) and MR CLEAN stroke trial (lower panel).

Although most trials collected outcome data at multiple time points, single time point analysis dominated. Furthermore, there was no consensus on the most appropriate timing of assessment within each field. For diseases with acute onset and monophasic course, a fixed time point analysis can miss the window of promising treatment effect. Longitudinal extensions to ordinal analysis have the potential to summarize the treatment effect over time better (Schildcrout et al., 2022). Alternatively, analyses can be repeated for multiple time points. For instance, in the latest trial of GBS (Walgaard et al., 2021), four separate common odds ratios were reported for GBS disability score at week 4, 8, 12, and 26 respectively. These might be pooled in a longitudinal proportional odds model to give an overall summary of treatment effect averaged from week 4 to week 26.

It is recommended to adjust for covariates that influence the prognosis of patients (Steyerberg et al., 2000). Statistically, it can increase power and protect against chance imbalances after randomization (Lingsma et al., 2010). Moreover, an unadjusted treatment effect is averaged over the trial population characteristics (comparison between the treated arm and the untreated arm) and might be less transferable to other populations compared to an adjusted treatment effect, which is conditionally on the same prognostic profiles of two patients (comparison between a treated patient and a similar untreated patient) (Austin et al., 2010). Practically, the choice of baseline characteristics to adjust for should be guided by subject matter knowledge (Hernández et al., 2005). We noted that many trials performed multiple subgroup analyses without correction for multiplicity. As noted often, subgroup analyses need to be interpreted cautiously, and mainly as exploratory analyses (Assmann et al., 2000; Hernández et al., 2006).

Our review has some limitations. The selected papers may not be fully representative of the general practice of neurological trials because we only focused on ordinal outcomes and five specific neurological diseases. The selection of studies in stroke was a convenient sample. We only included the most recent trials to prevent stroke trials from dominating the review, but many more trials have been conducted in this field. Methodological improvements have come especially from stroke and TBI research, such as the promotion of the PO model (Bath et al., 2007; Maas et al., 2010) and adjusting for baseline prognosis in the analysis (Collaboration, 2009; Hernández et al., 2004; Thompson et al., 2015).

Some studies dichotomized the ordinal scale but performed a time-to-event analysis (for instance, time to at least one point increase in GBS disability score). We counted such analysis as a dichotomization because the rank ordering information was lost, although a time-to-event analysis utilizes more information than a logistic regression analysis and can even provide more power than ordinal analysis under certain treatment effect patterns (Peterson et al., 2019). Finally, reporting was sometimes unclear on which method was adopted for the analysis of which outcome.

In conclusion, we identified directions of improvement for more efficient statistical analysis of ordinal outcomes in neurological trials:

- Exploit the ranking information contained in ordinal outcomes; provide visualizations to understand the full distribution of the ordinal scale between treatment arms.
- Adjust for important prognostic factors.
- Consider longitudinal analysis for treatment effect over time.

Further research needs to clarify the balance between maximizing the statistical power to detect a positive treatment effect and assumptions made in the statistical analysis.

## Supporting information

Data extraction item list

## Data Availability

All data produced in the present study are available upon reasonable request to the authors

## Contributorship statement

YL and SR contributed equally to this paper. YL: data extraction, statistical analysis, interpretation of results, writing of the manuscripts. SR: data extraction, statistical analysis, interpretation of results, writing of the manuscripts. LL: data extraction, interpretation of results, revision and edition of the manuscript. EW: interpretation of results, revision and edition of the manuscript. PD: interpretation of results, revision and edition of the manuscript. DD: interpretation of results, revision and edition of the manuscript. BJ: study conceptualization, methodology, revision and edition of the manuscript. ES: study conceptualization, methodology, revision and edition of the manuscript.

