## Supplementary figures and tables for "Opportunities for more powerful statistical analyses of ordinal scales: A scoping review of neurological trials"

### Search terms

**SAH**

("Subarachnoid Hemorrhage"[Mesh] OR "Subarachnoid Hemorrhage"[tw] OR "SAH"[tw] OR "Aneurysmal Subarachnoid Hemorrhage*"[tw] OR "Intracranial Subarachnoid Hemorrhage"[tw]) AND ((randomizedcontrolledtrial[Filter]) AND (2015:2021[pdat]))

**TBI**

("brain injuries, traumatic"[Mesh] OR "traumatic brain injur*"[tiab] OR "brain trauma*"[tiab] OR "TBI"[tiab] OR "traumatic encephalopath*"[tiab])

AND ((randomizedcontrolledtrial[Filter]) AND (2015:2021[pdat]))

**GBS**

("Guillain-Barre Syndrome"[Mesh] OR "Guillain-Barre Syndrome"[tw] OR "Guillain Barre Syndrome"[tw] OR "Guillaine-Barre Syndrome"[tw] OR "Guillaine Barre Syndrome"[tw] OR "Guillaine Barré Syndrome"[tw] OR "Guillain Barré Syndrome"[tw]) AND ((randomizedcontrolledtrial[Filter]) AND (2015:2021[pdat]))

**Stroke**

("Stroke"[Mesh] OR "Stroke*"[tiab] OR "Cerebrovascular Accident*"[tiab] OR "CVA"[tiab] OR "Brain Vascular Accident*"[tiab] OR "Acute Stroke*"[tiab] OR "Acute Cerebrovascular Accident"[tiab])AND ((randomizedcontrolledtrial[Filter]) AND (2015:2021[pdat]))

**Meningitis**

(“meningitis” [MeSH] OR “meningitis”[tw] OR “encephalitis”[tw]) AND ((randomizedcontrolledtrial[Filter]) AND (2015:2021[pdat]))

### Common ordinal outcome measure in neurology

| Scale | Scores |
| --- | --- |
| GBS DS (Guillain-Barré Syndrome Disability Score | 0 - A healthy state 1 - Minor symptoms and capable of running 2 - Able to walk 10m or more without assistance but unable to run  3 - Able to walk 10m across an open space with help 4 - Bedridden or chairbound 5 - Requiring assisted ventilation for at least part of the day 6 - Dead |
| GOS (Glasgow Outcome Scale) | 1 - Death  2 - Vegetative state  3 - Severe disability  4 - Moderate disability  5 - Low disability |
| GOS-E (Glasgow Outcome Scale-Extended) | 1 - Death  2 - Vegetative state  3 - Lower severe disability  4 - Upper severe disability  5 - Lower moderate disability  6 - Upper moderate disability  7 - Lower good recovery  8 - Upper good recovery |
| mRS (Modified Rankin Scale) | 0 - No symptoms 1 - No significant disability. Able to carry out all usual activities, despite some symptoms 2 - Slight disability. Able to look after own affairs without assistance, but unable to carry out all previous activities 3 - Moderate disability. Requires some help, but able to walk unassisted 4 - Moderately severe disability. Unable to attend to own bodily needs without assistance, and unable to walk unassisted 5 - Severe disability. Requires constant nursing care and attention, bedridden, incontinent 6 – Death |

### Supplementary figures and tables

**Table 1A.** Frequency of cut-off points for common ordinal measures in the 70 included studies

| Disease (n studies) | Most common ordinal outcome measure | Dichotomizations used for analysis of ordinal scale | Total frequency (how often used for analysis) |
| --- | --- | --- | --- |
| GBS (4) | GBS DS | 0-1 versus 2-6 | 1 |
|  |  | 0-2 versus 3-6 | 2 |
| Meningitis (7) | mRS | 0-1 versus 2-6 | 1 |
|  |  | 0-3 versus 4-6  0-4 versus 5-6 | 1  1 |
| SAH (10) | mRS | 0-2 versus 3-6 | 4 |
|  |  | 0-3 versus 4-6 | 2 |
|  | GOS | 1-3 versus 4-5 | 2 |
|  |  | 1-4 versus 5 | 2 |
|  | GOS-E | 1-4 versus 5-8  1-5 versus 6-8 | 1  1 |
| TBI (13) | GOS-E | 1-3 versus 4-8 | 1 |
|  |  | 1-4 versus 5-8 | 7 |
|  |  | Unknown | 1 |
| Stroke (36) | mRS | 0-1 versus 2-6 | 20 |
|  |  | 0-2 versus 3-6 | 27 |
|  |  | 0-3 versus 4-6 | 13 |
|  |  | 0-4 versus 5-6 | 5 |
|  |  | 0-5 versus 6 | 3 |

**Table 2A.** Frequency of time points selected for analysis for common ordinal measures in included 70 studies

| Disease (n studies) | Most common ordinal outcome measure | Time points of interest for analysis of ordinal scale (after start treatment) | Total frequency, % (how often used for analysis) |
| --- | --- | --- | --- |
| GBS (4) | GBS DS | 1 week  2 weeks  3 weeks  4 weeks  6 weeks  8 weeks  12 weeks  16 weeks  24 weeks  26 weeks  1 week after treatment completion  2 weeks after treatment completion | 1  1  1  3  1  2  2  1  1  1  1  1 |
| Meningitis (7) | mRS | 56 days  60 days  10 weeks  3 months  6 months  8 months  9 months  12 months | 1  1  1  1  2  1  1  1 |
| SAH (10) | mRS | At discharge  3 months  6 months  12 months | 2  3  3  1 |
|  | GOS | At discharge  3 months | 1  3 |
|  | GOS-E | At discharge  3 months  6 months | 1  1  1 |
| TBI (13) | GOS-E | At discharge  28 days  3 months  6 months  12 months  24 months | 1  1  3  9  2  2 |
| Stroke (36) | mRS | At discharge  14 days  1 month  30 – 43 days  90 days  6 months  24 months | 1  1  1  2  30  3  1 |

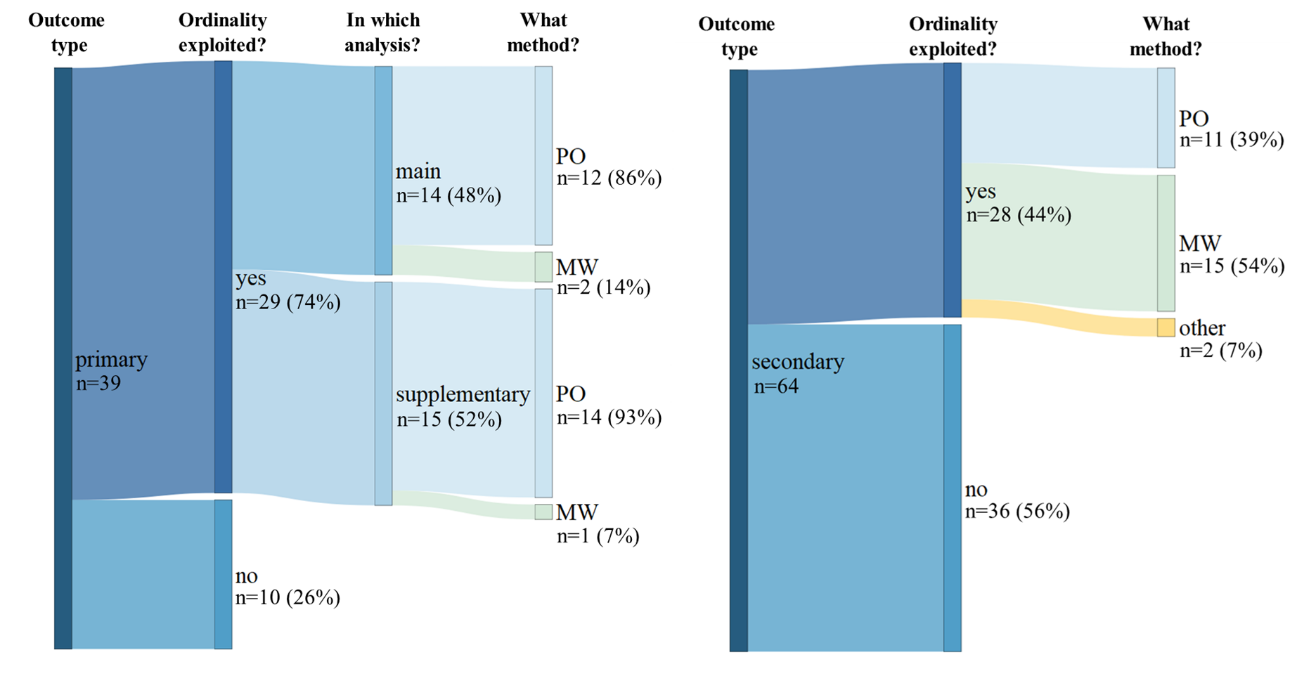

**Figure 1A.** Distribution of ordinal analysis and analysis methods for primary ordinal outcomes (left) and secondary ordinal outcomes (right). MW: Mann-Whitney U test or its associated variants such as van Elteren test, win odds, etc. PO: proportional odds model. Other: t-test, linear-by-linear association test.

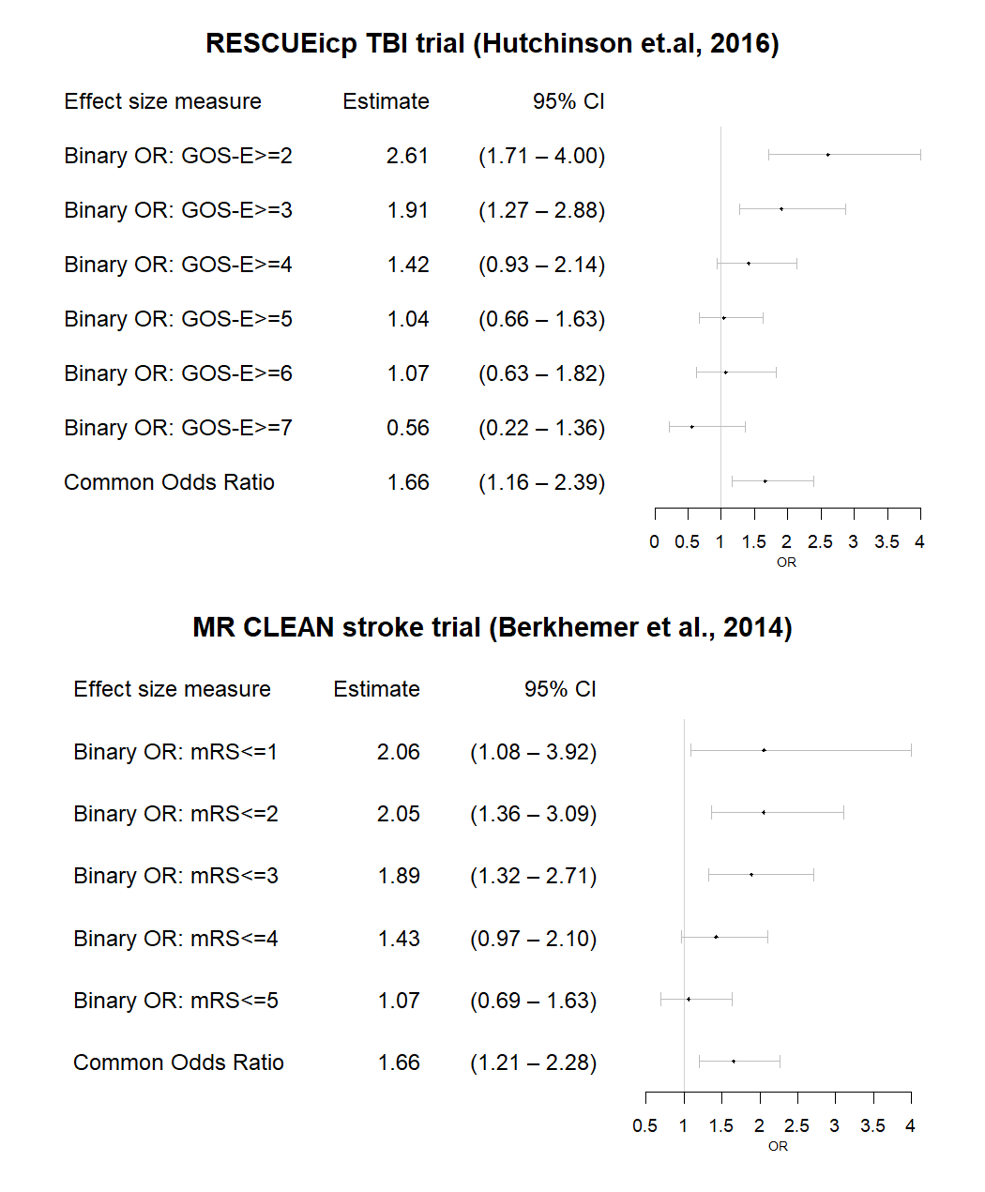

Figure 2A. Forest plots of odds ratios per cut-point from separate logistic regression models and the common odds ratio from the proportional odds model, for RESCUEicp TBI trial (upper panel) and MR CLEAN stroke trial (lower panel). The odds of a more favorable outcome condition (higher GOS-E score or lower mRS score) is modelled so a positive odds ratio means a beneficial effect.
