## Supplementary material for "Opportunities for more powerful statistical analyses of ordinal scales: A scoping review of neurological trials": Data extraction item list

### Items in the Data Extract Form used for screening and scoring trials

| Item extraction question | Possible answers |
| --- | --- |
| Minimal 1 ordinal outcome | Yes/no |
| Exploiting ordinality at least once | Yes/no |
| Dichotomizing (arbitrary cut-off only) | Yes/no |
| Dichotomizing as min ... grades change only | Yes/no |
| Dichotomizing only (both cut-off and min … grades increase/decrease) | Yes/no |
| Dichotomizing only (both cut-off and min … grades increase/decrease) | Yes/no |
| Treated as continuous only | Yes/no |
| Ordinal outcome is measured at multiple time points (after start treatment) | Yes/no |
| Ordinal outcome is measured at a single time point | Yes/no |
| Ordinal outcome is analyzed in a longitudinal way | Yes/no/NA(single time point) |
| Ordinal outcome is analyzed in separate analyses | Yes/no/NA(single time point) |
| Covariate adjustment in analysis of ordinal outcome | Yes/no |
| Subgroup analysis / stratification for analysis of ordinal outcome | Yes/no |
| How is the PO assumption reported | NA(no ordinal analysis)/not reported/reported, satisfied/reported, violated |
| Continuous outcome is measured at multiple time points (after start treatment) | Yes/no |
| Continuous outcome is measured at a single time point | Yes/no |
| Continuous outcome is analyzed in a longitudinal way | Yes/no/NA(single time point) |
| Continuous outcome is analyzed in separate analyses | Yes/no/NA(single time point) |
